# CohortSymmetry: An R package to perform sequence symmetry analysis using the OMOP common data model

**DOI:** 10.1101/2025.11.14.25340229

**Authors:** Xihang Chen, Ty Stanford, Yuchen Guo, Berta Raventós, Mike Du, Xintong Li, Amy Lam, George Corby, Núria Mercadé-Besora, Marta Alcalde-Herraiz, Kim López-Güell, Antonella Delmestri, Wai Yi Man, Daniel Prieto-Alhambra, Edward Burn, Martí Català, Nicole Pratt, Annika Jödicke, Danielle Newby

## Abstract

**Background:** Real-world data are valuable for detecting adverse drug events, and Sequence Symmetry Analysis (SSA) is a simple yet effective method frequently used for this purpose. However, heterogeneous implementations across studies limit reproducibility and scalability. To address this, we developed an open-source R package that standardises SSA analytics using data mapped to the Observational Medical Outcomes Partnership (OMOP) Common Data Model (CDM).

**Methods:** We developed *CohortSymmetry*, an R package that implements SSA for OMOP CDM data. The package was validated through unit testing and evaluated empirically by estimating adjusted sequence ratios (ASRs) with 95% confidence intervals (CIs) for 23 positive and 10 negative controls across six European databases, including CPRD GOLD (UK) and THIN® (Belgium, Italy, Romania, Spain, UK). Sensitivity and specificity were defined as the proportions of positive and negative controls correctly identified by SSA. Sensitivity analyses varied key parameters, including the washout period.

**Results:** *CohortSymmetry* passed high-coverage unit tests. Of 33 eligible controls, four showed results consistent with expectations across all databases; for example, the amiodarone–levothyroxine pair had a lower 95% CI bound >1 in each. Sensitivity was moderate, whereas specificity was high in the primary analyses. Parameter variation influenced outcomes; a 365-day prior observation requirement reduced specificity in CPRD GOLD from 75% to 38%.

**Conclusions:** *CohortSymmetry* enables reproducible SSA using OMOP CDM data. Differences across databases likely reflect heterogeneity in data capture and prescribing patterns. Limitations include residual data variability and SSA’s susceptibility to time-varying confounding, underscoring the need for tailored analytic design in pharmacovigilance studies.

**Key Messages:**

- We developed CohortSymmetry, an open-source R package that standardises SSA analytics using OMOP CDM-mapped data and verified the correctness of functions via unit testing and application to real-world datasets.
- CohortSymmetry passed high-coverage tests, and among 33 selected controls, four showed results consistent with expectations across all databases; varying analytical parameters affected results.
- The package provides a reproducible and scalable framework for multi-database SSA studies, supporting robust pharmacovigilance, but careful specification of parameters is required to account for the characteristics of the medical domain under investigation.

## Introduction

Adverse drug events (ADEs) are typically identified during clinical trials; however, limited follow-up durations and small sample sizes often prevent the detection of all ADEs before entering the market^1^. Furthermore, clinical trials are often restricted to specific populations and may exclude older adults, minority groups, or those with comorbidities^2,3^. This limits the generalisability of trial findings and highlights the importance of post-marketing surveillance. Real-world data (RWD), which capture routine clinical practice across broad and diverse populations, provide a valuable resource for monitoring drug safety in real-world settings.

In pharmacoepidemiology studies, Sequence Symmetry Analysis (SSA) is a type of self-controlled, case-only design used to assist with ADE detection^4^. SSA is computationally simple, requires fewer assumptions than many alternative designs, and has been used in contexts such as vaccine safety^5^ and allergy reactions^6^. Previous research has shown that SSA demonstrates moderate sensitivity and high specificity in detecting ADEs^7^. SSA assesses the asymmetry in the timing of initiation between two drugs or between a drug and a condition. A consistent pattern of one event following another may suggest that the first exposure causes an adverse effect, prompting treatment with the second drug or leading to a subsequent diagnosis^8^. In SSA, the drug under investigation is designated as the index drug, while the marker drug or condition, serves as a surrogate or direct indicator of the outcome.

The growing use of RWD in healthcare research^9^ has been accompanied by efforts to harmonise its structure through common data models (CDMs). These models enable the application of analytical approaches uniformly across diverse databases. Among them, the Observational Medical Outcomes Partnership (OMOP) CDM (https://www.ohdsi.org/data-standardization/) is widely adopted, offering a person-level framework that standardises both database organisation and clinical vocabularies^10^. However, no standardised analytical pipeline currently exists for conducting SSA for OMOP CDM mapped data. To address this gap, we developed CohortSymmetry^11^, an open-source R package that standardises SSA analytics. This paper describes its functionality and illustrates its use by applying it to a set of positive and negative controls identified from literature across European healthcare databases from Belgium, Italy, Romania, Spain and the UK.

## Methods

### Sequence Symmetry Analysis

SSA is a pharmacoepidemiological method frequently applied to investigate potential associations between a drug and an ADE^12^. It compares the number of individuals who experience the outcome of interest (marker event), typically as initiation of another drug or a diagnosis, after the initiation of the drug under investigation (index event) (*N*_*index → marker*_) with those who experienced it before (*N*_*marker → index*_). The crude sequence ratio (CSR) is then calculated as follows:

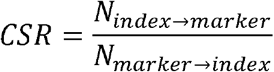

Under the null hypothesis of no association, the CSR should be close to 1. However, because prescribing or diagnosis trends may change over time (e.g., due to seasonality, the introduction of new drugs, or changes in diagnostic practices), the crude sequence ratio may falsely suggest an association between two events. To account for these temporal trends, the adjusted sequence ratio (ASR) was introduced. The ASR compares the observed sequence ratio to the ratio expected under a null scenario of no causal relationship, after adjusting for background changes in prescribing frequency over time^4^. This adjustment helps distinguish genuine temporal associations from those driven by underlying prescribing trends. Confidence intervals (CIs) for the ASR can be estimated using Jeffreys’ noninformative prior, a Bayesian approach that provides stable interval estimates even when event counts are small or data are sparse^13^.

### Package details and software dependencies

CohortSymmetry version 0.2.4 is on CRAN, written in R (4.2.3) and organised with roxygen^14^. Co-created with epidemiologists, CohortSymmetry is dependent on open-source R packages specifically developed for OMOP CDM including omock^15^, PatientProfiles^16^, omopgenerics^17^, DrugUtilisation^18^ and CodelistGenerator^19^. Comprehensive unit tests are implemented to ensure that the functions in CohortSymmetry package perform as intended^20^. These tests validate inputs, check core function logics, and verify the implementation of key parameters. Subject to these unit tests passing, CohortSymmetry has been refactored to optimise the run time and memory use and improve in-code documentation to enhance clarity and maintainability.

### Main functions

CohortSymmetry provides a comprehensive set of functions for calculating both crude and adjusted sequence ratios (CSRs and ASRs). See Table 1 for an overview of the main functions included in the package.

**Table 1.**
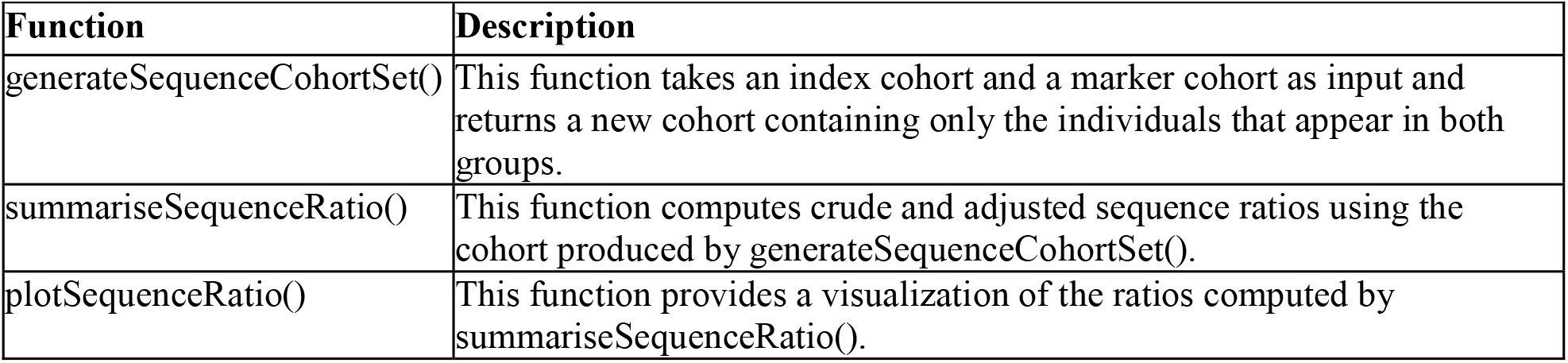
Main functions of the CohortSymmetry R package.

| Function | Description |
| --- | --- |
| generateSequenceCohortSet() | This function takes an index cohort and a marker cohort as input and returns a new cohort containing only the individuals that appear in both groups. |
| summariseSequenceRatio() | This function computes crude and adjusted sequence ratios using the cohort produced by generateSequenceCohortSet(). |
| plotSequenceRatio() | This function provides a visualization of the ratios computed by summariseSequenceRatio(). |

The first function to use is generateSequenceCohortSet(), which requires an index cohort and a marker cohort. Users can create these cohorts using OHDSI tools such as CohortConstructor^21^ or DrugUtilisation^18^. The function returns a cohort table containing individuals present in both the index and marker cohorts who meet the user-specified criteria. The function parameters require careful specification. For example, the cohortDateRange ensures at least one episode of both the index and marker occurs within the study period, which is why patient ID = 4 in Figure 1 was excluded. Further eligibility is then assessed based on the first observed episode of both the index and the marker cohort within cohortDateRange by specifying parameters including daysPriorObservation, washoutWindow, indexMarkerGap and combinationWindow. Detailed documentation and usage examples are available through the package vignettes (https://cran.r-project.org/web/packages/CohortSymmetry/vignettes/). Individuals who do not meet all specified criteria are excluded from the analysis.

**Figure 1.**
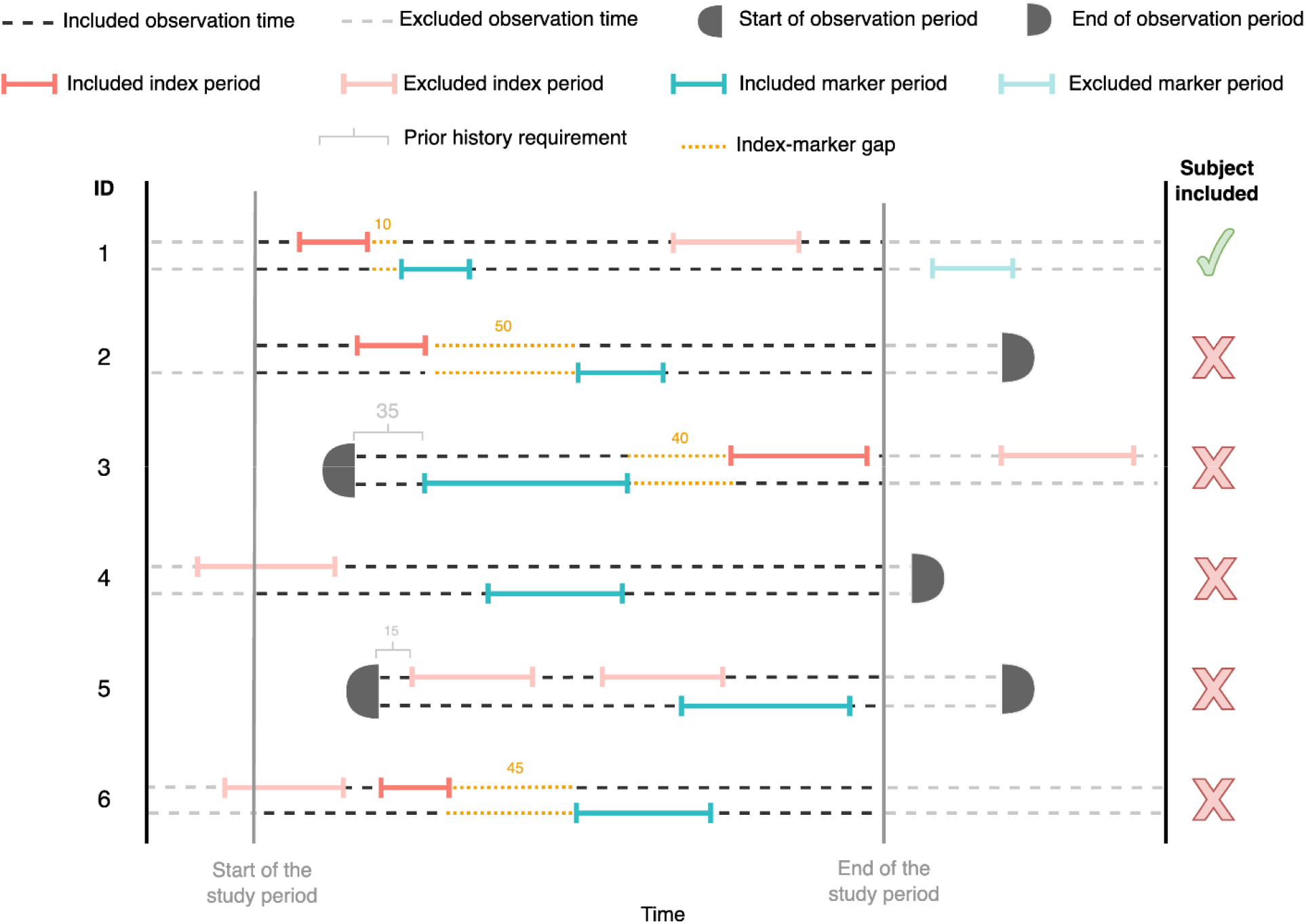
Illustration of how parameters interact in generateSequenceCohortSet(). Setting indexMarkerGap = 30 excludes patients with ID 2, 3, and 6. Setting cohortDateRange excludes patient with ID 4. Setting daysPriorObservation = 30 excludes patient with ID 5.

The cohort generated by generateSequenceCohortSet() is passed to the summariseSequenceRatio() function to calculate CSRs and ASRs. plotSequenceRatio() then generates forest plots of the sequence ratios with flexibility for the user to customise.

### Application of CohortSymmetry on European databases

A total of 23 positive and 10 negative controls were identified from the established pharmacoepidemiological literature and subsequently reviewed by clinical epidemiologists to confirm their classification. Positive controls were selected for their strong, previously documented associations, while negative controls were chosen due to a lack of biological plausibility or empirical evidence for a causal relationship. The full list and supporting references are provided in Supplementary Table S1^22–24^. The databases included were CPRD GOLD (UK) and the THIN® database (covering Belgium, Italy, Romania, Spain, and the UK), spanning the period from 1st January 2010 to 1st January 2021. While both are primary care databases, they differ significantly in their geographical and healthcare system coverage. CPRD GOLD provides a deep, nationally representative sample from the UK’s National Health Service (NHS). In contrast, THIN® offers a broader, multinational perspective, capturing data from primary care practices across different European healthcare systems. This combination allows the analysis to assess both the consistency of findings within a single, unified system (UK) and their generalizability across diverse European populations and clinical settings.

For the primary analyses, individuals were included if they had at least one episode of both the index and marker events, separated by an initiation gap of up to one year (combinationWindow = (0, 365)). A minimum of 365 days of prior observation was required, while no restrictions were applied to the washout window or the indexMarkerGap parameter. An example code of CohortSymmetry for the amiodarone-allopurinol pair can be found in Supplementary Table S2.

To evaluate the performance of the SSA method on the primary analyses, we calculated recall on control pairs which had enough counts^25,26^. From this, we calculated sensitivity and specificity, which are defined as the proportion of positive and negative control outcomes correctly identified by SSA, respectively (i.e., CI entirely > 1 and CI containing 1).

In sensitivity analyses, the washout window was set to 365 days, and the combinationWindow was extended to (0, 730) to evaluate the robustness of results. Results based on fewer than 50 exposed individuals were suppressed due to limited statistical power.

All analytical code is publicly available at: https://github.com/oxford-pharmacoepi/CohortSymmetryMethodsStudy.

## Results

### Package

CohortSymmetry is freely available under the Apache License (Version 2.0) and can be obtained from CRAN (https://cran.r-project.org/web/packages/CohortSymmetry/index.html), where full details and instructions for setup and use are provided. The source code underwent rigorous quality checks against PostgreSQL and SQL Server through the execution of more than 300 tests, obtaining over 95% test coverage.

### Application of CohortSymmetry on European databases

#### Primary Analyses

Out of 33 selected controls (23 positive control outcomes and 10 negative control outcomes), four had sufficient counts (Supplementary Table S3) and demonstrated the expected association across all six databases (Table 2). Of these, three were positive control outcomes (amiodarone-levothyroxine, NSAIDs-PPIs and opioids-constipation agents) and one was a negative control outcome (amiodarone-allopurinol). The positive control outcome bisphosphonates-PPIs showed a consistent association across all databases; however, the association was in the opposite direction than expected, as the upper limit of the 95% confidence interval was consistently above 1.

**Table 2.**
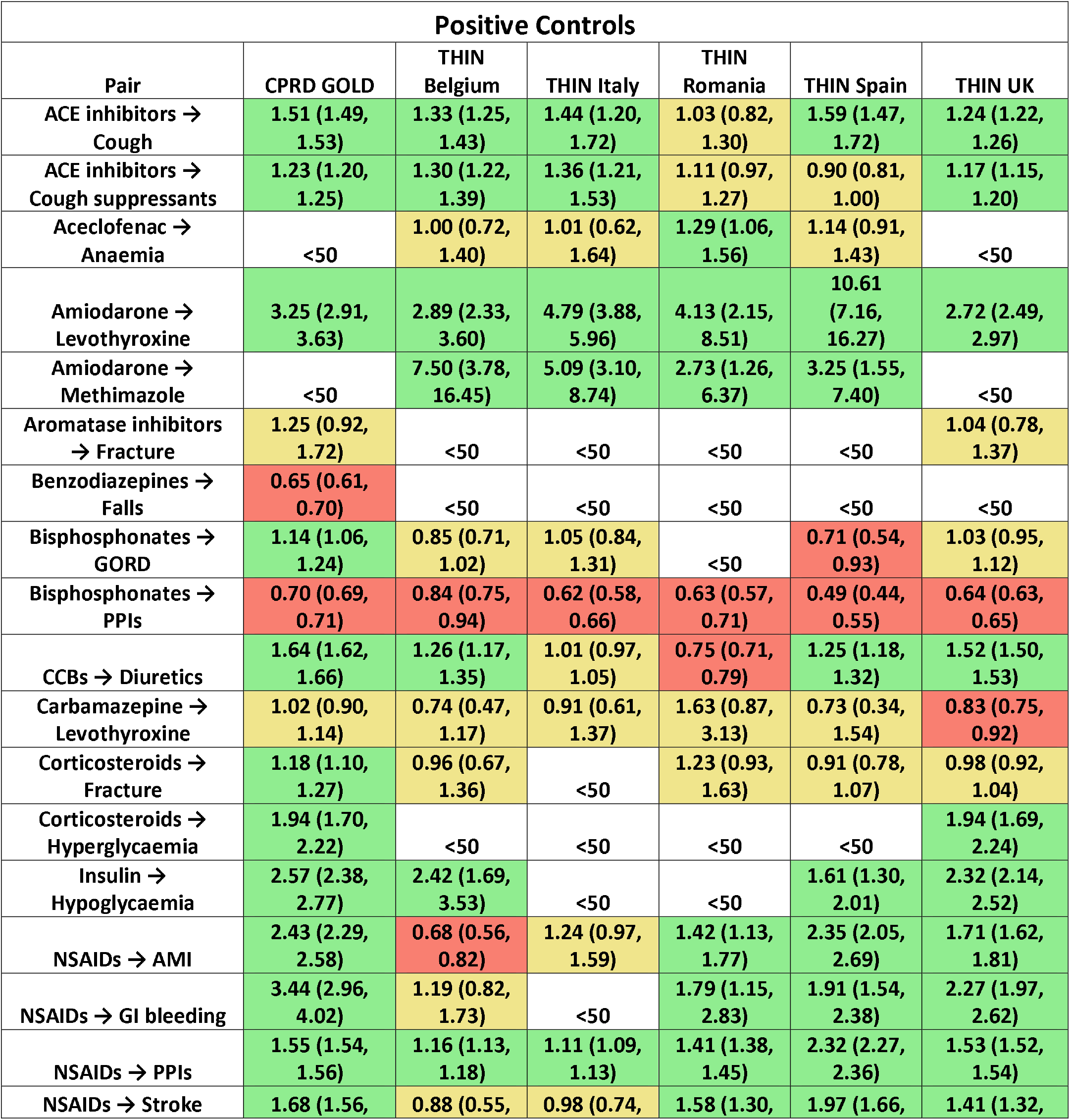
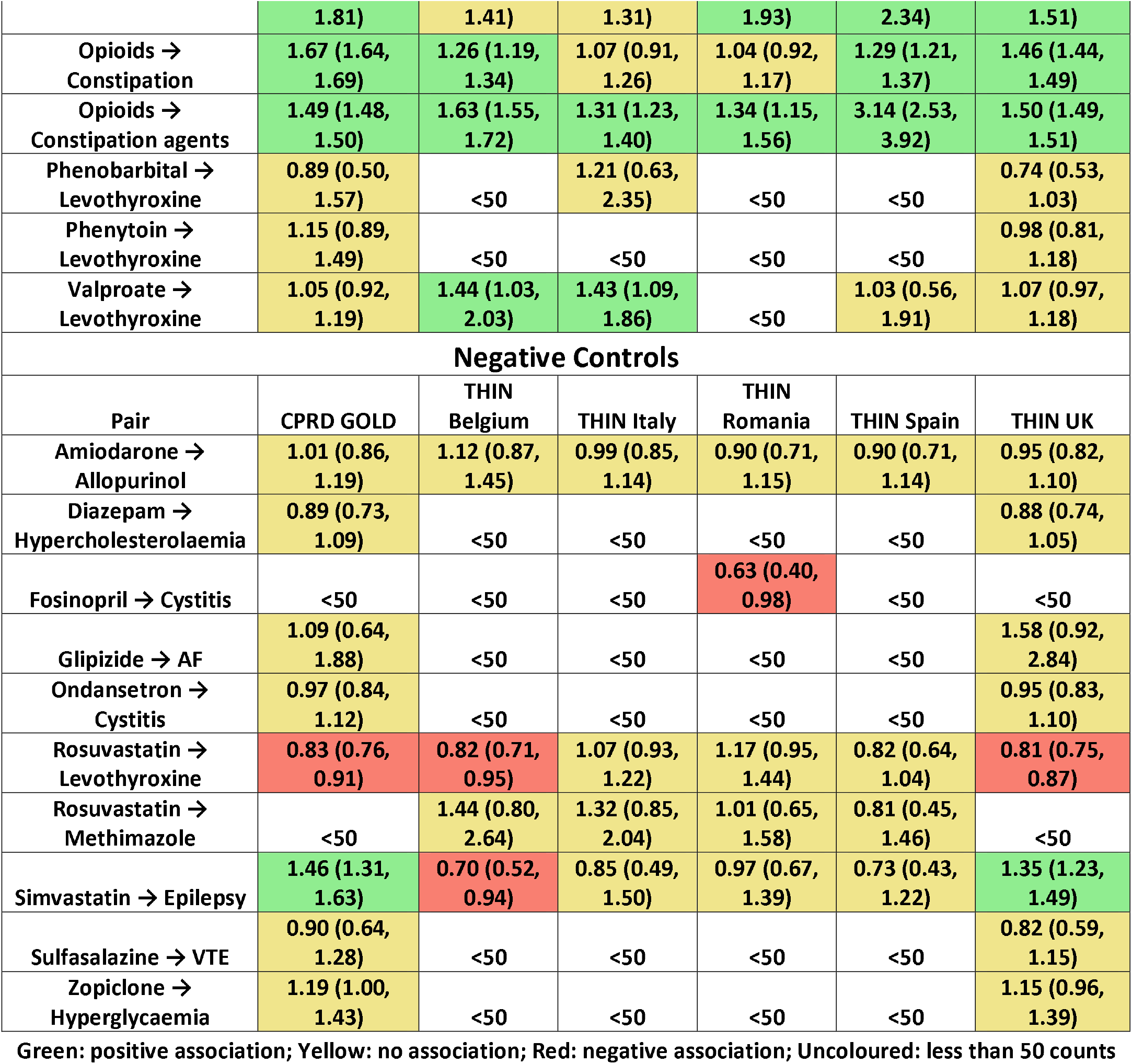
Primary analysis results – ASRs (with 95% CI) on the set of positive and negative controls. Control - outcome pairs with fewer than 50 events were excluded; ACE: Angiotensin-converting Enzyme; AF: Atrial Fibrillation; AMI: Acute myocardial infarction; CCB: Calcium channel blockers; GI: Gastro-intestinal; GORD: Gastro-oesophageal reflux disease; NSAID: Non-steroidal Anti-inflammatory Drug; PPI: Proton Pump Inhibitor; VTE: Venous Thromboembolism

The SSA demonstrated moderate sensitivity and specificity across most databases. For example, in CPRD GOLD, the database with the highest population count, SSA correctly identified 14 of 21 positive control outcomes with sufficient counts (sensitivity/recall = 67%) (Supplementary Figure S1, Supplementary Table S4). For negative control outcomes, it correctly produced the expected null association in 6 of 8 cases (specificity = 75%).

#### Sensitivity Analysis

When applying a 365-day prior washout requirement, four control outcomes continued to have sufficient counts and showed the expected associations across all six databases. These were the same controls identified in the main analysis. However, metrics such as specificity have been impacted. In the primary analysis, CPRD GOLD produced the expected null associations for 6 of 8 negative control outcomes, whereas under the washout requirement, this decreased to 3 of 8 (Supplementary Table S5).

When the parameter combinationWindow was set to 0-730 days, five pairs of control outcomes had sufficient counts and showed the expected associations across all six databases. Four of these were positive controls: amiodarone–levothyroxine, NSAIDs–PPIs, opioids-constipation, and opioids-constipation agents; and one was a negative control, amiodarone-allopurinol (Supplementary Table S6). When both washoutWindow was set to 365 days and combinationWindow to 0-730 days, again only these same five control outcomes had sufficient counts and showed the expected associations across all databases (Supplementary Table S7).

## Discussion

Identifying ADEs is critical for clinicians and policymakers involved in pharmacovigilance. To support this, we developed CohortSymmetry, an open-source R package that standardises the implementation of SSA for data mapped to the OMOP CDM. By reducing the need for customised code, CohortSymmetry improves reproducibility and accessibility for researchers with basic R experience. The package allows users to flexibly adjust parameters such as combinationWindow and washoutWindow to suit different study designs and clinical domains. We further evaluated the performance of CohortSymmetry using various positive and negative controls across multiple European databases and observed consistency in different signals, highlighting its utility and potential to be used in ADE signal detection across diverse healthcare settings.

CohortSymmetry was designed to align with established OHDSI tools such as CohortConstructor^21^ and DrugUtilisation^18^, facilitating its integration into broader observational research workflows. The package underwent extensive unit testing with high coverage, ensuring robust performance. However, current functionality is limited by the absence of built-in stratified analyses, preventing users from directly examining subpopulations by sex, age, or other characteristics. These can still be implemented with additional code using the OHDSI framework, and future versions of CohortSymmetry aim to incorporate native support for such analyses.

In this context, understanding the strengths and limitations of SSA is important for interpreting the role of CohortSymmetry in pharmacovigilance research. SSA is computationally efficient and straightforward to apply across large datasets, making it particularly suitable for early-stage signal detection. However, it remains susceptible to time-varying confounding and protopathic bias, meaning its outputs should be interpreted as hypothesis-generating rather than confirmatory^4^. CohortSymmetry therefore serves as a practical, standardised foundation for efficient ADE signal screening, serving as a hypothesis-generating framework that precedes more rigorous pharmacoepidemiological approaches, including propensity score-adjusted cohort analyses and their modern extensions^27–29^, self-controlled case series^30^, and instrumental variable methods^31^, which provide stronger causal inference beyond exploratory signal detection.

Using CohortSymmetry, we evaluated 23 positive and 10 negative control pairs across six European databases. Consistency in key signals demonstrated the tool’s reliability. For instance, the amiodarone-levothyroxine pair showed robust associations in all databases with sufficient counts, consistent with amiodarone’s known thyroid effects that often necessitate levothyroxine treatment^32^. The adjusted sequence ratios (ASRs) ranged from 2.46 to 4.96, similar to values reported in an Asia-Pacific SSA study (1.52-5.30)^22^. Likewise, the amiodarone-methimazole pair consistently showed a positive association in non-UK databases but not in UK datasets, likely reflecting local prescribing preferences where carbimazole is used instead of methimazole^33^. An interesting observation was the bisphosphonate-PPI pair, which exhibited a negative association across databases despite being a positive control. This may reflect clinicians’ preventive prescribing behaviour of using PPIs to mitigate bisphosphonate-induced gastrointestinal irritation - an effect also observed in other SSA studies^34^. Variability in ASRs across databases can be partly explained by differences in prescribing patterns and population demographics. For example, simvastatin–epilepsy ASRs ranged from 0.36 to 1.53, possibly due to variation in statin use across countries, with simvastatin prescribed more frequently in the UK than in Italy or Spain^35^. Additionally, differences in population demographics, such as age and sex composition, may further explain the variation in ASRs across countries.

This study demonstrated high specificity and moderate sensitivity in the primary analysis, consistent with previous literature^22^. The specificity values observed in this study were lower than the 91% reported by Pratt *et al*., likely due to differences in the databases and control outcomes used. Parameter settings, particularly prior observation requirements, substantially influenced specificity, underscoring the need to tailor SSA parameters to specific contexts.

The study’s strengths include its multi-database design, which enhanced generalisability and enabled assessment of cross-country consistency, as well as the systematic variation of analytic parameters to evaluate robustness across settings. Using multiple European databases allowed us to examine both methodological reliability and real-world heterogeneity in drug utilisation, thereby strengthening the external validity of our findings. Nonetheless, several limitations should be noted. Aside from the methodological limitations inherent to SSA, one limitation is the overlap between the two UK databases (CPRD GOLD and THIN®)^36,37^ may have introduced partial duplication of patient populations, potentially affecting cross-database comparisons.

## Conclusion

CohortSymmetry is effective for performing SSA studies within the OMOP CDM framework, as supported by unit testing and the analyses conducted in this study. Testing a set of controls across various European databases demonstrated that several controls were robust across diverse datasets and resilient to changes in key parameters. Differences observed between European databases likely stem from variations in data sources and prescribing practices. The clinical findings also highlight the importance of tailoring analytical settings to the clinical context and characteristics of each dataset to ensure reliable signal detection.

## Data Availability

All data produced in the present work are contained in the manuscript.

## List of Abbreviations

ACE: Angiotensin-converting Enzyme
AE: Adverse Event
AF: Atrial Fibrillation
ASR: Adjusted Sequence Ratio
CI: Confidence Interval
CSR: Crude Sequence Ratio
FDR: False discovery rate
GORD: Gastro-oesophageal reflux disease
NSAID: Non-steroidal Anti-inflammatory Drug
OHDSI: Observational Health Data Sciences and Informatics
OMOP CDM: Observational Medical Outcomes Partnership Common Data Model
PPI: Proton Pump Inhibitor
RWD: Real-world Data
SSA: Sequence Symmetry Analysis
VTE: Venous Thromboembolism

## Acknowledgement

None.

## Notes

**Funding information** The author(s) declare that financial support was received for the research, authorship, and/or publication of this work. This research was partially funded by the European Health Data and Evidence Network (EHDEN). This activity under the EHDEN has received funding from the Innovative Medicines Initiative 2 (IMI2) Joint Undertaking under grant agreement No 806968. IMI2 receives support from the European Union’s Horizon 2020 research and innovation program and European Federation of Pharmaceutical Industries and Associations (EFPIA). The sponsors of the study did not have any involvement in the writing of the manuscript or the decision to submit it for publication. Additionally, there was partial support from the Oxford NIHR Biomedical Research Centre.

**Conflict of interest statement** Professor Daniel Prieto-Alhambra research group has received research fundings from the European Medicines Agency and Innovative Medicines Initiative; and grants from Amgen, Chiesi-Taylor, Gilead, Lilly, Janssen, Novartis, and UCB Biopharma; and consultancy or speaker fees (paid to the University) from UCB Biopharma. Moreover, Janssen has funded or supported training programs organized by DPA’s department. BR works for a research group that receives/received unconditional research grants from UCB, Johnson and Johnson, Innovative Medicines Initiative, and the European Medicines Agency, none of which relate the content of this manuscript.

### Competing Interest Statement

The authors have declared no competing interest.

### Funding Statement

The author(s) declare that financial support was received for the research, authorship, and/or publication of this work. This research was partially funded by the European Health Data and Evidence Network (EHDEN). This activity under the EHDEN has received funding from the Innovative Medicines Initiative 2 (IMI2) Joint Undertaking under grant agreement No 806968. IMI2 receives support from the European Union's Horizon 2020 research and innovation program and European Federation of Pharmaceutical Industries and Associations (EFPIA). The sponsors of the study did not have any involvement in the writing of the manuscript or the decision to submit it for publication. Additionally, there was partial support from the Oxford NIHR Biomedical Research Centre.

### Author Declarations

The use of CPRD GOLD for this study was approved through CPRD's Research Data Governance (RDG) Process (Protocol number: 22_002351). Individual consent was not necessary, as the CPRD GOLD data are de-identified and approved for research use by the UK Health Research Authority and the NHS Health and Social Care Research Ethics Committee. The use of the THIN databases for this study was approved by THIN EuroBoard under protocol 2024_10 and provided by Cegedim. The data are available from Cegedim upon request for researchers who meet the criteria for access to confidential data. THIN databases contain de‐identified data provided by patients as part of their routine care; therefore, no informed consent was required as well.

