## Supplementary Material for "CohortSymmetry: An R package to perform sequence symmetry analysis using the OMOP common data model"

### Supplementary Table S1: List of positive and negative controls identified from literature

| Index event | Marker event | Type | Control | Reference (DOI or Link) |
| --- | --- | --- | --- | --- |
| Bisphosphonates  (ATC) | GORD | Drug-Condition | Positive | National Institute for Health and Care Excellence. BNF: British National Formulary. <https://bnf.nice.org.uk/>  Accessed 22 July 2025  10.1038/s41597-022-01159-y |
| NSAIDs (ATC) | GI bleeding | Drug-Condition | Positive | National Institute for Health and Care Excellence. BNF: British National Formulary. <https://bnf.nice.org.uk/>  Accessed 22 July 2025  10.1038/s41597-022-01159-y |
| NSAIDs (ATC) | AMI | Drug-Condition | Positive | National Institute for Health and Care Excellence. BNF: British National Formulary. <https://bnf.nice.org.uk/>  Accessed 22 July 2025  10.1038/s41597-022-01159-y |
| ACE inhibitors (ATC) | Cough | Drug-Condition | Positive | National Institute for Health and Care Excellence. BNF: British National Formulary. <https://bnf.nice.org.uk/>  Accessed 22 July 2025  10.1038/s41597-022-01159-y |
| Aceclofenac (ingredient) | Anaemia | Drug-Condition | Positive | National Institute for Health and Care Excellence. BNF: British National Formulary. <https://bnf.nice.org.uk/>  Accessed 22 July 2025  10.1038/s41597-022-01159-y |
| Opioids (ATC) | Constipation | Drug-Condition | Positive | National Institute for Health and Care Excellence. BNF: British National Formulary. <https://bnf.nice.org.uk/>  Accessed 22 July 2025  10.1038/s41597-022-01159-y |
| Aromatase inhibitors (ATC) | Fracture | Drug-Condition | Positive | National Institute for Health and Care Excellence. BNF: British National Formulary. <https://bnf.nice.org.uk/>  Accessed 22 July 2025  10.1038/s41597-022-01159-y |
| Benzodiazepines  (ATC) | Falls | Drug-Condition | Positive | National Institute for Health and Care Excellence. BNF: British National Formulary. <https://bnf.nice.org.uk/>  Accessed 22 July 2025  10.1038/s41597-022-01159-y |
| Corticosteroids (ATC) | Fracture | Drug-Condition | Positive | National Institute for Health and Care Excellence. BNF: British National Formulary. <https://bnf.nice.org.uk/>  Accessed 22 July 2025  10.1038/s41597-022-01159-y |
| NSAIDs (ATC) | Stroke | Drug-Condition | Positive | National Institute for Health and Care Excellence. BNF: British National Formulary. <https://bnf.nice.org.uk/>  Accessed 22 July 2025  10.1038/s41597-022-01159-y |
| Insulin (ATC) | Hypoglycaemia | Drug-Condition | Positive | National Institute for Health and Care Excellence. BNF: British National Formulary. <https://bnf.nice.org.uk/>  Accessed 22 July 2025  10.1038/s41597-022-01159-y |
| Corticosteroids (ATC) | Hyperglycaemia | Drug-Condition | Positive | National Institute for Health and Care Excellence. BNF: British National Formulary. <https://bnf.nice.org.uk/>  Accessed 22 July 2025  10.1038/s41597-022-01159-y |
| Bisphosphonates (ATC) | PPIs (ATC) | Drug-Drug | Positive | National Institute for Health and Care Excellence. BNF: British National Formulary. <https://bnf.nice.org.uk/>  Accessed 22 July 2025  10.1038/s41597-022-01159-y  PPIs are a drug proxy for “GORD” |
| NSAIDs (ATC) | PPIs (ATC) | Drug-Drug | Positive | National Institute for Health and Care Excellence. BNF: British National Formulary. <https://bnf.nice.org.uk/>  Accessed 22 July 2025  10.1038/s41597-022-01159-y  PPIs are a drug proxy for “GI bleeding” |
| Opioids (ATC) | Constipation Agents (ATC) | Drug-Drug | Positive | National Institute for Health and Care Excellence. BNF: British National Formulary. <https://bnf.nice.org.uk/>  Accessed 22 July 2025  10.1038/s41597-022-01159-y  Constipation agents are drug proxies for “constipation” |
| Amiodarone (ingredient) | Levothyroxine (ingredient) | Drug-Drug | Positive | National Institute for Health and Care Excellence. BNF: British National Formulary. <https://bnf.nice.org.uk/>  Accessed 22 July 2025  [10.1002/pds.3780](https://doi.org/10.1002/pds.3780)  Levothyroxine is a drug proxy for “hypothyroidism” |
| Amiodarone (ingredient) | Methimazole (ingredient) | Drug-Drug | Positive | National Institute for Health and Care Excellence. BNF: British National Formulary. <https://bnf.nice.org.uk/>  Accessed 22 July 2025  10.1038/s41597-022-01159-y  Methimazole is a drug proxy for “hypothyroidism” |
| Carbamazepine (ingredient) | Levothyroxine (ingredient) | Drug-Drug | Positive | https://pubmed.ncbi.nlm.nih.gov/23946049/ |
| Phenobarbital (ingredient) | Levothyroxine (ingredient) | Drug-Drug | Positive | https://pubmed.ncbi.nlm.nih.gov/23946049/ |
| Phenytoin (ingredient) | Levothyroxine (ingredient) | Drug-Drug | Positive | https://pubmed.ncbi.nlm.nih.gov/23946049/ |
| Valproate (ingredient) | Levothyroxine (ingredient) | Drug-Drug | Positive | National Institute for Health and Care Excellence. BNF: British National Formulary. <https://bnf.nice.org.uk/>  Accessed 22 July 2025  10.1038/s41597-022-01159-y  Valproate is a drug proxy for “hypothyroidism” |
| CCBs (ATC) | Diuretics (ATC) | Drug-Drug | Positive | National Institute for Health and Care Excellence. BNF: British National Formulary. <https://bnf.nice.org.uk/>  Accessed 22 July 2025  10.1038/s41597-022-01159-y  Diuretics are a drug proxy for “oedema” |
| ACE inhibitors (ATC) | Antitussives (ATC) | Drug-Drug | Positive | National Institute for Health and Care Excellence. BNF: British National Formulary. <https://bnf.nice.org.uk/>  Accessed 22 July 2025  10.1038/s41597-022-01159-y  Antitussives are a drug proxy for “cough” |
| Amiodarone (ingredient) | Allopurinol (ingredient) | Drug-Drug | Negative | [10.1002/pds.3780](https://doi.org/10.1002/pds.3780) |
| Rosuvastatin (ingredient) | Levothyroxine (ingredient) | Drug-Drug | Negative | 10.1038/s41597-022-01159-y |
| Rosuvastatin (ingredient) | Methimazole (ingredient) | Drug-Drug | Negative | 10.1038/s41597-022-01159-y |
| Glipizide (ingredient) | AF | Drug-Condition | Negative | 10.1038/s41597-022-01159-y |
| Fosinopril (ingredient) | Cystitis | Drug-Condition | Negative | 10.1038/s41597-022-01159-y |
| Ondansetron (ingredient) | Cystitis | Drug-Condition | Negative | 10.1038/s41597-022-01159-y |
| Simvastatin (ingredient) | Epilepsy | Drug-Condition | Negative | 10.1038/s41597-022-01159-y |
| Diazepam (ingredient) | High cholesterol | Drug-Condition | Negative | 10.1038/s41597-022-01159-y |
| Zopiclone (ingredient) | Hyperglycaemia | Drug-Condition | Negative | 10.1038/s41597-022-01159-y |
| Sulfasalazine (ingredient) | VTE | Drug-Condition | Negative | 10.1038/s41597-022-01159-y |

ACE: Angiotensin-converting Enzyme; AF: Atrial Fibrillation; AMI: Acute myocardial infarction; CCB: Calcium channel blockers; GI: Gastro-intestinal; GORD: Gastro-oesophageal reflux disease; NSAID: Non-steroidal Anti-inflammatory Drug; PPI: Proton Pump Inhibitor; VTE: Venous Thromboembolism

### Supplementary Table S2: Example usage of the CohortSymmetry R package

| **Example** | **R code** |
| --- | --- |
| Given that the cohorts amiodarone and allopurinol are present in the cdm reference, this step gives another cohort in the cdm named amiodarone_allopurinol. | cdm <- CohortSymmetry::generateSequenceCohortSet(  cdm = cdm,  indexTable = "amiodarone",  markerTable = "allopurinol",  name = "amiodarone_allopurinol",  cohortDateRange = as.Date(c("2010-01-01", "2021-12-31")),  daysPriorObservation = 365,  washoutWindow = 0,  indexMarkerGap = Inf,  combinationWindow = c(0, 365),  movingAverageRestriction = 548  ) |
| Using the cohort named amiodarone_allopurinol, this gives the desired sequence ratios together with 95% CIs. | sequence_ratios <- CohortSymmetry::summariseSequenceRatios(  cohort = cdm$amiodarone_allopurinol,  confidenceInterval = 95  ) |
| Using the R object sequence_ratios produced from the last step, this step provides forest plots to visualise the results. | plotSequenceRatios(  result = sequence_ratios,  onlyASR = FALSE,  plotTitle = "Sequence Ratios Forest Plots for Amiodarone - Allopurinol",  labs = c("Sequence Ratios", "Control Pairs"),  colours = c("red", "blue")  ) |

### Supplementary Table S3: Counts of each control for the main analysis

| **Positive Controls** | | | | | | |
| --- | --- | --- | --- | --- | --- | --- |
| **Pair** | **CPRD GOLD** | **THIN Belgium** | **THIN Italy** | **THIN Romania** | **THIN Spain** | **THIN UK** |
| **ACE inhibitors → Cough** | **45958** | **6414** | **1797** | **1896** | **1912** | **49478** |
| **ACE inhibitors → Cough suppressants** | **112252** | **5826** | **697** | **667** | **3866** | **69403** |
| **Aceclofenac → Anaemia** | **<5** | **178** | **52** | **667** | **397** | **<5** |
| **Amiodarone → Levothyroxine** | **1521** | **271** | **643** | **<5** | **201** | **2039** |
| **Amiodarone → Methimazole** | **2030** | **660** | **989** | **68** | **479** | **2500** |
| **Aromatase inhibitors → Fracture** | **<5** | **77** | **133** | **0** | **0** | **<5** |
| **Benzodiazepines → Falls** | **165** | **<5** | **<5** | **<5** | **<5** | **273** |
| **Bisphosphonates → GORD** | **2783** | **927** | **451** | **<5** | **524** | **3428** |
| **Bisphosphonates → PPIs** | **62238** | **2234** | **7746** | **3654** | **5472** | **65574** |
| **CCBs → Diuretics** | **136796** | **6194** | **20929** | **18459** | **12057** | **159943** |
| **Carbamazepine → Levothyroxine** | **1498** | **145** | **236** | **0** | **0** | **1898** |
| **Corticosteroids → Fracture** | **3641** | **170** | **<5** | **305** | **718** | **5430** |
| **Corticosteroids → Hyperglycaemia** | **1039** | **<5** | **<5** | **<5** | **<5** | **995** |
| **Insulin → Hypoglycaemia** | **4335** | **245** | **<5** | **<5** | **548** | **5206** |
| **NSAIDs → AMI** | **6166** | **1133** | **506** | **736** | **1886** | **8101** |
| **NSAIDs → GI bleeding** | **1124** | **238** | **<5** | **172** | **548** | **1274** |
| **NSAIDs → PPIs** | **415936** | **54790** | **112403** | **72053** | **106938** | **413858** |
| **NSAIDs → Stroke** | **3547** | **189** | **334** | **1090** | **1064** | **4578** |
| **Opioids → Constipation** | **93299** | **7564** | **743** | **1573** | **5528** | **93283** |
| **Opioids → Constipation agents** | **374532** | **8894** | **4750** | **1034** | **555** | **342367** |
| **Phenobarbital → Levothyroxine** | **0** | **<5** | **93** | **<5** | **<5** | **165** |
| **Phenytoin → Levothyroxine** | **324** | **<5** | **<5** | **<5** | **<5** | **509** |
| **Valproate → Levothyroxine** | **1521** | **271** | **643** | **<5** | **201** | **2039** |
| **Negative Controls** | | | | | | |
| **Amiodarone → Allopurinol** | **668** | **389** | **1251** | **385** | **647** | **755** |
| **Diazepam → Hypercholesterolaemia** | **490** | **<5** | **<5** | **<5** | **<5** | **780** |
| **Fosinopril → Cystitis** | **<5** | **<5** | **<5** | **208** | **<5** | **<5** |
| **Glipizide → AF** | **0** | **<5** | **<5** | **<5** | **<5** | **71** |
| **Rosuvastatin → Levothyroxine** | **2426** | **1434** | **2153** | **728** | **925** | **3370** |
| **Rosuvastatin → Methimazole** | **<5** | **0** | **162** | **146** | **0** | **<5** |
| **Simvastatin → Epilepsy** | **1995** | **471** | **68** | **309** | **186** | **3662** |
| **Sulfasalazine → VTE** | **148** | **<5** | **<5** | **<5** | **<5** | **195** |
| **Zopiclone → Hyperglycaemia** | **546** | **<5** | **<5** | **<5** | **<5** | **550** |

### Supplementary Table S4: Metric recall of positive and negative controls per database

| Databases | Control Type | N | Recall |
| --- | --- | --- | --- |
| CPRD GOLD | Positive | 21 | 67% |
| CPRD GOLD | Negative | 8 | 75% |
| THIN Belgium | Positive | 18 | 56% |
| THIN Belgium | Negative | 4 | 50% |
| THIN Italy | Positive | 16 | 44% |
| THIN Italy | Negative | 4 | 80% |
| THIN Romania | Positive | 15 | 53% |
| THIN Romania | Negative | 5 | 80% |
| THIN Spain | Positive | 18 | 61% |
| THIN Spain | Negative | 4 | 100% |
| THIN UK | Positive | 20 | 60% |
| THIN UK | Negative | 8 | 75% |

### Supplementary Table S5: Complete results of ASRs with 95% CI of the sensitivity analysis (365 days of washout)

| **Positive Controls** | | | | | | |
| --- | --- | --- | --- | --- | --- | --- |
| **Pair** | **CPRD GOLD** | **THIN Belgium** | **THIN Italy** | **THIN Romania** | **THIN Spain** | **THIN UK** |
| **ACE inhibitors → Cough** | **1.45 (1.43, 1.47)** | **1.33 (1.25, 1.43)** | **1.44 (1.20, 1.72)** | **1.03 (0.82, 1.30)** | **1.59 (1.47, 1.72)** | **1.24 (1.22, 1.26)** |
| **ACE inhibitors → Cough suppressants** | **1.23 (1.21, 1.26)** | **1.30 (1.22, 1.39)** | **1.36 (1.21, 1.53)** | **1.11 (0.97, 1.27)** | **0.90 (0.81, 1.00)** | **1.17 (1.15, 1.20)** |
| **Aceclofenac → Anaemia** | **<50** | **1.00 (0.72, 1.40)** | **1.01 (0.62, 1.64)** | **1.29 (1.06, 1.56)** | **1.14 (0.91, 1.43)** | **<50** |
| **Amiodarone → Levothyroxine** | **3.60 (3.20, 4.06)** | **2.89 (2.33, 3.60)** | **4.79 (3.88, 5.96)** | **4.13 (2.15, 8.51)** | **10.61 (7.16, 16.27)** | **2.72 (2.49, 2.97)** |
| **Amiodarone → Methimazole** | **<50** | **7.50 (3.78, 16.45)** | **5.09 (3.10, 8.74)** | **2.73 (1.26, 6.37)** | **3.25 (1.55, 7.40)** | **<50** |
| **Aromatase inhibitors → Fracture** | **1.17 (0.85, 1.61)** | **<50** | **<50** | **<50** | **<50** | **1.04 (0.78, 1.37)** |
| **Benzodiazepines → Falls** | **0.63 (0.59, 0.68)** | **<50** | **<50** | **<50** | **<50** | **<50** |
| **Bisphosphonates → GORD** | **1.05 (0.97, 1.14)** | **0.85 (0.71, 1.02)** | **1.05 (0.84, 1.31)** | **<50** | **0.71 (0.54, 0.93)** | **1.03 (0.95, 1.12)** |
| **Bisphosphonates → PPIs** | **0.63 (0.62, 0.65)** | **0.84 (0.75, 0.94)** | **0.62 (0.58, 0.66)** | **0.63 (0.57, 0.71)** | **0.49 (0.44, 0.55)** | **0.64 (0.63, 0.65)** |
| **CCBs → Diuretics** | **1.77 (1.75, 1.80)** | **1.26 (1.17, 1.35)** | **1.01 (0.97, 1.05)** | **0.75 (0.71, 0.79)** | **1.25 (1.18, 1.32)** | **1.52 (1.50, 1.53)** |
| **Carbamazepine → Levothyroxine** | **1.07 (0.94, 1.22)** | **0.74 (0.47, 1.17)** | **0.91 (0.61, 1.37)** | **1.63 (0.87, 3.13)** | **0.73 (0.34, 1.54)** | **0.83 (0.75, 0.92)** |
| **Corticosteroids → Fracture** | **1.12 (1.04, 1.20)** | **0.96 (0.67, 1.36)** | **<50** | **1.23 (0.93, 1.63)** | **0.91 (0.78, 1.07)** | **0.98 (0.92, 1.04)** |
| **Corticosteroids → Hyperglycaemia** | **1.88 (1.64, 2.16)** | **<50** | **<50** | **<50** | **<50** | **1.94 (1.69, 2.24)** |
| **Insulin → Hypoglycaemia** | **1.90 (1.74, 2.08)** | **2.42 (1.69, 3.53)** | **<50** | **<50** | **1.61 (1.30, 2.01)** | **2.32 (2.14, 2.52)** |
| **NSAIDs → AMI** | **2.24 (2.11, 2.38)** | **0.68 (0.56, 0.82)** | **1.24 (0.97, 1.59)** | **1.42 (1.13, 1.77)** | **2.35 (2.05, 2.69)** | **1.71 (1.62, 1.81)** |
| **NSAIDs → GI bleeding** | **3.20 (2.74, 3.74)** | **1.19 (0.82, 1.73)** | **<50** | **1.79 (1.15, 2.83)** | **1.91 (1.54, 2.38)** | **2.27 (1.97, 2.62)** |
| **NSAIDs → PPIs** | **1.64 (1.63, 1.65)** | **1.16 (1.13, 1.18)** | **1.11 (1.09, 1.13)** | **1.41 (1.38, 1.45)** | **2.32 (2.27, 2.36)** | **1.53 (1.52, 1.54)** |
| **NSAIDs → Stroke** | **1.56 (1.45, 1.68)** | **0.88 (0.55, 1.41)** | **0.98 (0.74, 1.31)** | **1.58 (1.30, 1.93)** | **1.97 (1.66, 2.34)** | **1.41 (1.32, 1.51)** |
| **Opioids → Constipation** | **1.49 (1.47, 1.51)** | **1.26 (1.19, 1.34)** | **1.07 (0.91, 1.26)** | **1.04 (0.92, 1.17)** | **1.29 (1.21, 1.37)** | **1.46 (1.44, 1.49)** |
| **Opioids → Constipation agents** | **1.55 (1.53, 1.56)** | **1.63 (1.55, 1.72)** | **1.31 (1.23, 1.40)** | **1.34 (1.15, 1.56)** | **3.14 (2.53, 3.92)** | **1.50 (1.49, 1.51)** |
| **Phenobarbital → Levothyroxine** | **0.71 (0.35, 1.41)** | **<50** | **1.21 (0.63, 2.35)** | **<50** | **<50** | **0.74 (0.53, 1.03)** |
| **Phenytoin → Levothyroxine** | **1.16 (0.87, 1.56)** | **<50** | **<50** | **<50** | **<50** | **0.98 (0.81, 1.18)** |
| **Valproate → Levothyroxine** | **1.02 (0.88, 1.17)** | **1.44 (1.03, 2.03)** | **1.43 (1.09, 1.86)** | **<50** | **1.03 (0.56, 1.91)** | **1.07 (0.97, 1.18)** |
| **Negative Controls** | | | | | | |
| **Pair** | **CPRD GOLD** | **THIN Belgium** | **THIN Italy** | **THIN Romania** | **THIN Spain** | **THIN UK** |
| **Amiodarone → Allopurinol** | **1.06 (0.90, 1.26)** | **1.12 (0.87, 1.45)** | **0.99 (0.85, 1.14)** | **0.90 (0.71, 1.15)** | **0.90 (0.71, 1.14)** | **0.95 (0.82, 1.10)** |
| **Diazepam → Hypercholesterolaemia** | **0.89 (0.73, 1.09)** | **<50** | **<50** | **<50** | **<50** | **0.88 (0.74, 1.05)** |
| **Fosinopril → Cystitis** | **<50** | **<50** | **<50** | **0.63 (0.40, 0.98)** | **<50** | **<50** |
| **Glipizide → AF** | **0.99 (0.56, 1.77)** | **<50** | **<50** | **<50** | **<50** | **1.58 (0.92, 2.84)** |
| **Ondansetron → Cystitis** | **0.94 (0.82, 1.09)** | **<50** | **<50** | **<50** | **<50** | **0.95 (0.83, 1.10)** |
| **Rosuvastatin → Levothyroxine** | **0.84 (0.76, 0.93)** | **0.82 (0.71, 0.95)** | **1.07 (0.93, 1.22)** | **1.17 (0.95, 1.44)** | **0.82 (0.64, 1.04)** | **0.81 (0.75, 0.87)** |
| **Rosuvastatin → Methimazole** | **<50** | **1.44 (0.80, 2.64)** | **1.32 (0.85, 2.04)** | **1.01 (0.65, 1.58)** | **0.81 (0.45, 1.46)** | **<50** |
| **Simvastatin → Epilepsy** | **1.39 (1.24, 1.56)** | **0.70 (0.52, 0.94)** | **0.85 (0.49, 1.50)** | **0.97 (0.67, 1.39)** | **0.73 (0.43, 1.22)** | **1.35 (1.23, 1.49)** |
| **Sulfasalazine → VTE** | **0.78 (0.54, 1.12)** | **<50** | **<50** | **<50** | **<50** | **0.82 (0.59, 1.15)** |
| **Zopiclone → Hyperglycaemia** | **1.18 (0.98, 1.41)** | **<50** | **<50** | **<50** | **<50** | **1.15 (0.96, 1.39)** |

### Supplementary Table S6: Complete results of ASRs with 95% CI of the sensitivity analysis (combination window of between 0 and 730 days)

| **Positive Controls** | | | | | | |
| --- | --- | --- | --- | --- | --- | --- |
| **Pair** | **CPRD GOLD** | **THIN Belgium** | **THIN Italy** | **THIN Romania** | **THIN Spain** | **THIN UK** |
| **ACE inhibitors → Cough** | **1.40 (1.38, 1.41)** | **1.24 (1.18, 1.31)** | **1.35 (1.17, 1.56)** | **1.13 (0.94, 1.36)** | **1.25 (1.17, 1.33)** | **1.33 (1.31, 1.34)** |
| **ACE inhibitors → Cough suppressants** | **1.28 (1.26, 1.30)** | **1.18 (1.12, 1.24)** | **1.22 (1.12, 1.33)** | **1.12 (1.01, 1.25)** | **0.85 (0.79, 0.93)** | **1.35 (1.33, 1.37)** |
| **Aceclofenac → Anaemia** | **<50** | **1.05 (0.80, 1.38)** | **1.07 (0.73, 1.56)** | **1.25 (1.08, 1.45)** | **1.12 (0.95, 1.33)** | **<50** |
| **Amiodarone → Levothyroxine** | **2.72 (2.49, 2.98)** | **2.74 (2.29, 3.29)** | **4.39 (3.68, 5.28)** | **4.38 (2.69, 7.44)** | **7.88 (5.86, 10.77)** | **2.58 (2.39, 2.78)** |
| **Amiodarone → Methimazole** | **<50** | **7.62 (4.22, 14.80)** | **5.95 (3.96, 9.21)** | **4.15 (2.22, 8.28)** | **5.47 (2.92, 11.07)** | **<50** |
| **Aromatase inhibitors → Fracture** | **1.16 (0.92, 1.48)** | **<50** | **<50** | **<50** | **<50** | **1.06 (0.85, 1.32)** |
| **Benzodiazepines → Falls** | **0.75 (0.70, 0.79)** | **<50** | **<50** | **<50** | **<50** | **<50** |
| **Bisphosphonates → GORD** | **1.16 (1.09, 1.23)** | **0.87 (0.75, 1.00)** | **1.13 (0.95, 1.34)** | **<50** | **0.81 (0.66, 0.99)** | **1.03 (0.97, 1.10)** |
| **Bisphosphonates → PPIs** | **0.72 (0.71, 0.73)** | **0.80 (0.73, 0.88)** | **0.61 (0.58, 0.64)** | **0.60 (0.55, 0.65)** | **0.45 (0.41, 0.50)** | **0.65 (0.64, 0.66)** |
| **CCBs → Diuretics** | **1.52 (1.50, 1.53)** | **1.29 (1.22, 1.36)** | **1.02 (0.99, 1.06)** | **0.70 (0.67, 0.73)** | **1.20 (1.14, 1.25)** | **1.46 (1.45, 1.47)** |
| **Carbamazepine → Levothyroxine** | **1.05 (0.96, 1.15)** | **0.62 (0.42, 0.90)** | **0.87 (0.63, 1.18)** | **1.16 (0.73, 1.84)** | **0.79 (0.43, 1.43)** | **0.73 (0.67, 0.79)** |
| **Corticosteroids → Fracture** | **1.24 (1.18, 1.31)** | **0.89 (0.67, 1.17)** | **<50** | **0.97 (0.80, 1.17)** | **1.02 (0.91, 1.15)** | **1.06 (1.01, 1.11)** |
| **Corticosteroids → Hyperglycaemia** | **1.88 (1.69, 2.10)** | **<50** | **<50** | **<50** | **<50** | **1.82 (1.63, 2.03)** |
| **Insulin → Hypoglycaemia** | **3.25 (3.05, 3.47)** | **2.82 (2.08, 3.89)** | **<50** | **<50** | **1.98 (1.65, 2.38)** | **2.89 (2.70, 3.10)** |
| **NSAIDs → AMI** | **2.60 (2.49, 2.71)** | **0.89 (0.77, 1.03)** | **1.39 (1.15, 1.69)** | **1.71 (1.44, 2.03)** | **2.61 (2.35, 2.89)** | **1.88 (1.81, 1.96)** |
| **NSAIDs → GI bleeding** | **3.49 (3.10, 3.94)** | **1.71 (1.27, 2.31)** | **<50** | **2.11 (1.49, 3.03)** | **1.85 (1.57, 2.18)** | **2.43 (2.17, 2.72)** |
| **NSAIDs → PPIs** | **1.54 (1.53, 1.55)** | **1.25 (1.23, 1.28)** | **1.17 (1.15, 1.19)** | **1.49 (1.46, 1.52)** | **2.40 (2.37, 2.44)** | **1.50 (1.49, 1.51)** |
| **NSAIDs → Stroke** | **2.08 (1.97, 2.21)** | **1.37 (0.98, 1.93)** | **1.22 (0.98, 1.52)** | **1.82 (1.57, 2.12)** | **2.26 (1.98, 2.58)** | **1.72 (1.63, 1.82)** |
| **Opioids → Constipation** | **1.76 (1.74, 1.78)** | **1.29 (1.24, 1.35)** | **1.18 (1.03, 1.34)** | **1.10 (1.01, 1.21)** | **1.33 (1.27, 1.40)** | **1.56 (1.54, 1.57)** |
| **Opioids → Constipation agents** | **1.49 (1.48, 1.50)** | **1.62 (1.55, 1.69)** | **1.33 (1.26, 1.40)** | **1.33 (1.18, 1.49)** | **3.04 (2.55, 3.65)** | **1.51 (1.51, 1.52)** |
| **Phenobarbital → Levothyroxine** | **0.97 (0.62, 1.50)** | **<50** | **0.82 (0.50, 1.37)** | **<50** | **<50** | **0.87 (0.65, 1.17)** |
| **Phenytoin → Levothyroxine** | **1.13 (0.92, 1.39)** | **<50** | **<50** | **<50** | **<50** | **0.96 (0.82, 1.13)** |
| **Valproate → Levothyroxine** | **1.03 (0.93, 1.14)** | **1.26 (0.95, 1.66)** | **1.18 (0.96, 1.46)** | **<50** | **1.13 (0.71, 1.81)** | **0.99 (0.91, 1.07)** |
| **Negative Controls** | | | | | | |
| **Pair** | **CPRD GOLD** | **THIN Belgium** | **THIN Italy** | **THIN Romania** | **THIN Spain** | **THIN UK** |
| **Amiodarone → Allopurinol** | **1.01 (0.89, 1.14)** | **0.96 (0.78, 1.18)** | **1.02 (0.90, 1.15)** | **0.87 (0.72, 1.05)** | **0.96 (0.79, 1.15)** | **0.97 (0.86, 1.09)** |
| **Diazepam → Hypercholesterolaemia** | **1.02 (0.88, 1.18)** | **<50** | **<50** | **<50** | **<50** | **0.91 (0.80, 1.03)** |
| **Fosinopril → Cystitis** | **<50** | **<50** | **<50** | **0.64 (0.46, 0.89)** | **<50** | **<50** |
| **Glipizide → AF** | **1.30 (0.89, 1.92)** | **<50** | **<50** | **<50** | **<50** | **1.65 (1.07, 2.62)** |
| **Ondansetron → Cystitis** | **0.82 (0.73, 0.92)** | **<50** | **<50** | **<50** | **<50** | **0.82 (0.74, 0.92)** |
| **Rosuvastatin → Levothyroxine** | **0.87 (0.81, 0.93)** | **0.73 (0.64, 0.82)** | **0.91 (0.81, 1.02)** | **1.12 (0.96, 1.31)** | **0.75 (0.61, 0.90)** | **0.73 (0.68, 0.78)** |
| **Rosuvastatin → Methimazole** | **<50** | **1.04 (0.66, 1.63)** | **1.01 (0.70, 1.45)** | **1.14 (0.81, 1.62)** | **0.73 (0.47, 1.16)** | **<50** |
| **Simvastatin → Epilepsy** | **1.59 (1.45, 1.73)** | **0.68 (0.54, 0.86)** | **1.43 (0.90, 2.31)** | **1.15 (0.86, 1.54)** | **0.84 (0.54, 1.29)** | **1.51 (1.39, 1.64)** |
| **Sulfasalazine → VTE** | **1.32 (1.02, 1.73)** | **<50** | **<50** | **<50** | **<50** | **1.01 (0.78, 1.31)** |
| **Zopiclone → Hyperglycaemia** | **1.22 (1.06, 1.41)** | **<50** | **<50** | **<50** | **<50** | **1.15 (0.99, 1.34)** |

### Supplementary Table S7: Complete results of ASRs with 95% CI of the sensitivity analysis (washout of 365 days and combination window of between 0 and 730 days)

| **Positive Controls** | | | | | | |
| --- | --- | --- | --- | --- | --- | --- |
| **Pair** | **CPRD GOLD** | **THIN Belgium** | **THIN Italy** | **THIN Romania** | **THIN Spain** | **THIN UK** |
| **ACE inhibitors → Cough** | **1.30 (1.29, 1.32)** | **1.24 (1.18, 1.31)** | **1.35 (1.17, 1.56)** | **1.13 (0.94, 1.36)** | **1.25 (1.17, 1.33)** | **1.33 (1.31, 1.34)** |
| **ACE inhibitors → Cough suppressants** | **1.28 (1.26, 1.30)** | **1.18 (1.12, 1.24)** | **1.22 (1.12, 1.33)** | **1.12 (1.01, 1.25)** | **0.85 (0.79, 0.93)** | **1.35 (1.33, 1.37)** |
| **Aceclofenac → Anaemia** | **<5** | **1.05 (0.80, 1.38)** | **1.07 (0.73, 1.56)** | **1.25 (1.08, 1.45)** | **1.12 (0.95, 1.33)** | **<5** |
| **Amiodarone → Levothyroxine** | **2.97 (2.70, 3.27)** | **2.74 (2.29, 3.29)** | **4.39 (3.68, 5.28)** | **4.38 (2.69, 7.44)** | **7.88 (5.86, 10.77)** | **2.58 (2.39, 2.78)** |
| **Amiodarone → Methimazole** | **<5** | **7.62 (4.22, 14.80)** | **5.95 (3.96, 9.21)** | **4.15 (2.22, 8.28)** | **5.47 (2.92, 11.07)** | **<5** |
| **Aromatase inhibitors → Fracture** | **1.07 (0.84, 1.37)** | **<5** | **<5** | **<5** | **<5** | **1.06 (0.85, 1.32)** |
| **Benzodiazepines → Falls** | **0.71 (0.67, 0.76)** | **<5** | **<5** | **<5** | **<5** | **<5** |
| **Bisphosphonates → GORD** | **1.04 (0.97, 1.10)** | **0.87 (0.75, 1.00)** | **1.13 (0.95, 1.34)** | **<5** | **0.81 (0.66, 0.99)** | **1.03 (0.97, 1.10)** |
| **Bisphosphonates → PPIs** | **0.66 (0.65, 0.67)** | **0.80 (0.73, 0.88)** | **0.61 (0.58, 0.64)** | **0.60 (0.55, 0.65)** | **0.45 (0.41, 0.50)** | **0.65 (0.64, 0.66)** |
| **CCBs → Diuretics** | **1.63 (1.62, 1.65)** | **1.29 (1.22, 1.36)** | **1.02 (0.99, 1.06)** | **0.70 (0.67, 0.73)** | **1.20 (1.14, 1.25)** | **1.46 (1.45, 1.47)** |
| **Carbamazepine → Levothyroxine** | **1.08 (0.98, 1.20)** | **0.62 (0.42, 0.90)** | **0.87 (0.63, 1.18)** | **1.16 (0.73, 1.84)** | **0.79 (0.43, 1.43)** | **0.73 (0.67, 0.79)** |
| **Corticosteroids → Fracture** | **1.16 (1.10, 1.22)** | **0.89 (0.67, 1.17)** | **<5** | **0.97 (0.80, 1.17)** | **1.02 (0.91, 1.15)** | **1.06 (1.01, 1.11)** |
| **Corticosteroids → Hyperglycaemia** | **1.76 (1.58, 1.97)** | **<5** | **<5** | **<5** | **<5** | **1.82 (1.63, 2.03)** |
| **Insulin → Hypoglycaemia** | **2.15 (2.00, 2.32)** | **2.82 (2.08, 3.89)** | **<5** | **<5** | **1.98 (1.65, 2.38)** | **2.89 (2.70, 3.10)** |
| **NSAIDs → AMI** | **2.33 (2.23, 2.44)** | **0.89 (0.77, 1.03)** | **1.39 (1.15, 1.69)** | **1.71 (1.44, 2.03)** | **2.61 (2.35, 2.89)** | **1.88 (1.81, 1.96)** |
| **NSAIDs → GI bleeding** | **3.18 (2.82, 3.60)** | **1.71 (1.27, 2.31)** | **<5** | **2.11 (1.49, 3.03)** | **1.85 (1.57, 2.18)** | **2.43 (2.17, 2.72)** |
| **NSAIDs → PPIs** | **1.59 (1.58, 1.60)** | **1.25 (1.23, 1.28)** | **1.17 (1.15, 1.19)** | **1.49 (1.46, 1.52)** | **2.40 (2.37, 2.44)** | **1.50 (1.49, 1.51)** |
| **NSAIDs → Stroke** | **1.90 (1.79, 2.01)** | **1.37 (0.98, 1.93)** | **1.22 (0.98, 1.52)** | **1.82 (1.57, 2.12)** | **2.26 (1.98, 2.58)** | **1.72 (1.63, 1.82)** |
| **Opioids → Constipation** | **1.50 (1.48, 1.52)** | **1.29 (1.24, 1.35)** | **1.18 (1.03, 1.34)** | **1.10 (1.01, 1.21)** | **1.33 (1.27, 1.40)** | **1.56 (1.54, 1.57)** |
| **Opioids → Constipation agents** | **1.50 (1.49, 1.51)** | **1.62 (1.55, 1.69)** | **1.33 (1.26, 1.40)** | **1.33 (1.18, 1.49)** | **3.04 (2.55, 3.65)** | **1.51 (1.51, 1.52)** |
| **Phenobarbital → Levothyroxine** | **0.68 (0.40, 1.15)** | **<5** | **0.82 (0.50, 1.37)** | **<5** | **<5** | **0.87 (0.65, 1.17)** |
| **Phenytoin → Levothyroxine** | **1.12 (0.89, 1.42)** | **<5** | **<5** | **<5** | **<5** | **0.96 (0.82, 1.13)** |
| **Valproate → Levothyroxine** | **0.98 (0.88, 1.10)** | **1.26 (0.95, 1.66)** | **1.18 (0.96, 1.46)** | **<5** | **1.13 (0.71, 1.81)** | **0.99 (0.91, 1.07)** |
| **Negative Controls** | | | | | | |
| **Pair** | **CPRD GOLD** | **THIN Belgium** | **THIN Italy** | **THIN Romania** | **THIN Spain** | **THIN UK** |
| **Amiodarone → Allopurinol** | **1.08 (0.95, 1.23)** | **0.96 (0.78, 1.18)** | **1.02 (0.90, 1.15)** | **0.87 (0.72, 1.05)** | **0.96 (0.79, 1.15)** | **0.97 (0.86, 1.09)** |
| **Diazepam → Hypercholesterolaemia** | **1.02 (0.88, 1.18)** | **<5** | **<5** | **<5** | **<5** | **0.91 (0.80, 1.03)** |
| **Fosinopril → Cystitis** | **<5** | **<5** | **<5** | **0.64 (0.46, 0.89)** | **<5** | **<5** |
| **Glipizide → AF** | **1.24 (0.83, 1.85)** | **<5** | **<5** | **<5** | **<5** | **1.65 (1.07, 2.62)** |
| **Ondansetron → Cystitis** | **0.80 (0.72, 0.90)** | **<5** | **<5** | **<5** | **<5** | **0.82 (0.74, 0.92)** |
| **Rosuvastatin → Levothyroxine** | **0.88 (0.81, 0.95)** | **0.73 (0.64, 0.82)** | **0.91 (0.81, 1.02)** | **1.12 (0.96, 1.31)** | **0.75 (0.61, 0.90)** | **0.73 (0.68, 0.78)** |
| **Rosuvastatin → Methimazole** | **<5** | **1.04 (0.66, 1.63)** | **1.01 (0.70, 1.45)** | **1.14 (0.81, 1.62)** | **0.73 (0.47, 1.16)** | **<5** |
| **Simvastatin → Epilepsy** | **1.47 (1.34, 1.61)** | **0.68 (0.54, 0.86)** | **1.43 (0.90, 2.31)** | **1.15 (0.86, 1.54)** | **0.84 (0.54, 1.29)** | **1.51 (1.39, 1.64)** |
| **Sulfasalazine → VTE** | **1.14 (0.86, 1.50)** | **<5** | **<5** | **<5** | **<5** | **1.01 (0.78, 1.31)** |
| **Zopiclone → Hyperglycaemia** | **1.18 (1.02, 1.36)** | **<5** | **<5** | **<5** | **<5** | **1.15 (0.99, 1.34)** |

### Supplementary Figure S1: Forest plot of point estimates and 95% confidence intervals for positive and negative control outcomes in the CPRD GOLD database

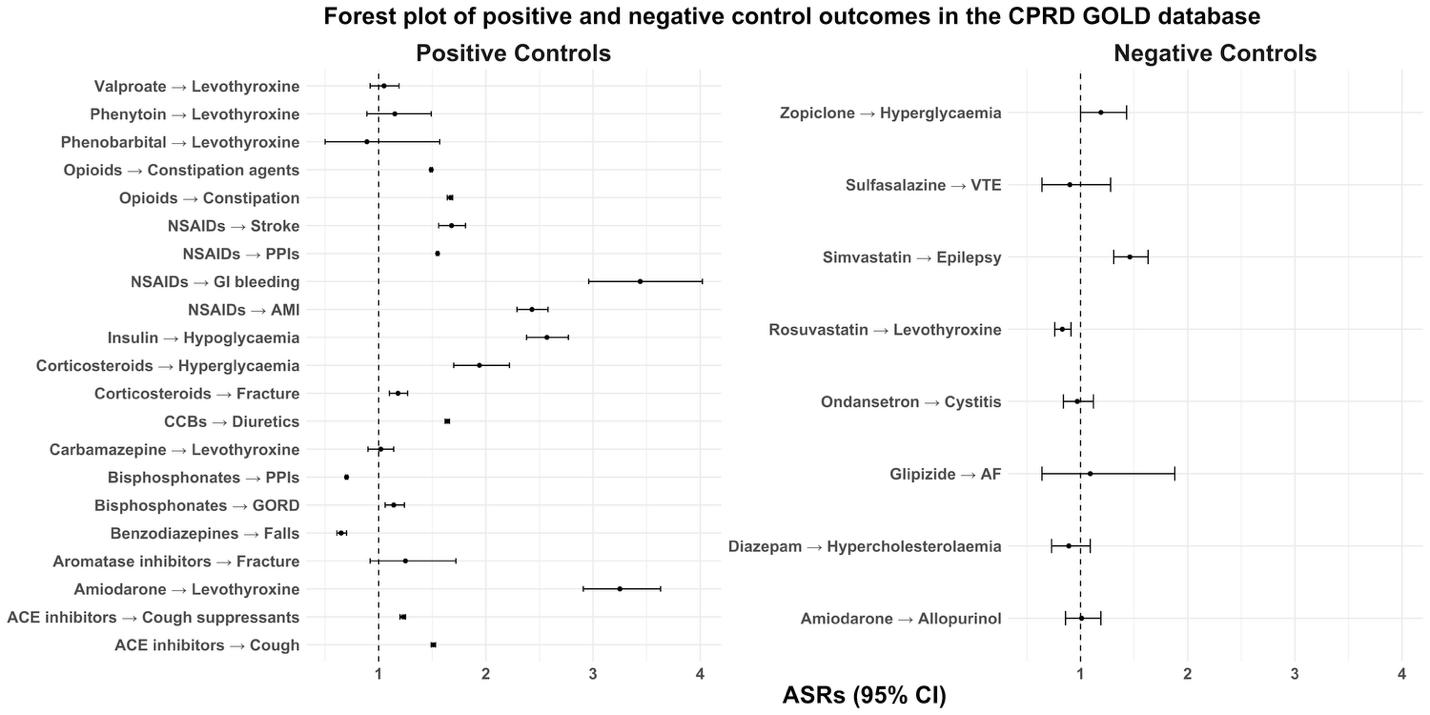
